# Randomized controlled trial combining constraint-induced movement therapy and action-observation training in unilateral cerebral palsy: clinical effects and influencing factors of treatment response

**DOI:** 10.1101/19009472

**Authors:** Cristina Simon-Martinez, Lisa Mailleux, Jasmine Hoskens, Els Ortibus, Ellen Jaspers, Nicole Wenderoth, Giuseppina Sgandurra, Giovanni Cioni, Guy Molenaers, Katrijn Klingels, Hilde Feys

## Abstract

**Introduction:** Constraint-induced movement therapy (CIMT) improves upper limb (UL) motor execution in unilateral cerebral palsy (uCP). As these children also show motor planning deficits, action-observation training (AOT) might be of additional value. Here, we investigated the combined effect of AOT to CIMT and identified factors influencing treatment response.

**Methods:** Forty-four children with uCP (mean 9y6m, SD 1y10m) participated in a 9-day camp wearing a splint for 6 hours/day and were allocated to the CIMT+AOT (n=22) and the CIMT+placebo group (n=22). The CIMT+AOT group received 15 hours of AOT (i.e. video-observation) and executed the observed tasks, whilst the CIMT+AOT group watched videos free of biological motion and executed the same tasks. The primary outcome measure was bimanual performance. Secondary outcomes included measures at body function and activity level assessed before (T1), after (T2) the intervention, and at 6 months follow-up (T3). Influencing factors included *behavioural* and *neurological* characteristics.

**Results:** Although no between-groups differences were found (p>0.05), the addition of AOT led to higher gains in children with initially poorer bimanual performance (p=0.02). Both groups improved in all outcome measures after the intervention and retained the gains at follow up (p<0.01). Poor sensory function resulted in larger improvements in the total group (p=0.03) and high amount of mirror movements tended to result in better response to the additional AOT training (p=0.06). Improvements were similar irrespective of the type of brain lesion or corticospinal tract wiring pattern.

**Conclusions:** Adding AOT to CIMT, resulted in better outcome for children with poor motor function and high amount of mirror movements. CIMT with or without AOT seems to be more beneficial for children with poor sensory function.

**Trial registration:** Registered at ClinicalTrials.gov on 22^nd^ August 2017 (Identifier: NCT03256357).

## 1. Introduction

The quality of life of children with unilateral cerebral palsy (uCP) can be compromised by their upper limb (UL) sensorimotor problems [1,2]. In the last decades, improving these deficits has been the focus of many studies. Constraint Induced Movement Therapy (CIMT) is one of the few treatments that has been thoroughly investigated and proven to be effective [3]. It consists of constraining the less impaired hand while intensively training the more impaired UL to promote increased use of the latter [4,5]. Whilst CIMT focuses on improving UL movement quality and efficiency, children with uCP also show deficits in motor representation and motor planning [6,7]. To overcome these deficits, Action-Observation Training (AOT) has been suggested as a potential treatment modality. AOT is based on the well-established principle that observation of actions activates the same cortical structures that are active during the actual performance of the task [8]. Although preliminary evidence has shown positive effects of AOT on UL function in children with uCP [9–13], it remains unknown whether the combination of CIMT and AOT would enhance the effect of CIMT on improving UL sensorimotor function in these children.

Despite increasing evidence proving the effectiveness of CIMT, the large variability in reported results leads to overall small to medium effect sizes [14,15]. Therefore, identifying factors influencing treatment response would contribute to the development of more efficient and more individualized treatment planning. Poor initial hand function has influenced a better response to CIMT in previous studies [16,17], but there remains controversy regarding age [16–18]. In adult stroke survivors, there is preliminary evidence that sensory deficits may also influenced UL motor outcome [19,20]. We hypothesize that children with impaired sensory function might also benefit more from the intensive use of their more impaired hand. Similarly, mirror movements have a negative impact on UL function [21–23], yet their value to influence treatment response remains unknown. Among potential neurological factors are the underlying lesion type (predominantly white matter vs. grey matter damage) and the corticospinal tract (CST) wiring pattern, due to their value in explaining variability in UL function [24–26]. Thus far, only one small study showed improvements after CIMT regardless of the lesion type [27]. However, there is controversy regarding the influence of the CST wiring pattern on treatment response [27–29]. Some studies have reported a worse outcome after CIMT in children with an ipsilateral CST wiring [28–30] whilst others showed positive outcome irrespective of the CST wiring pattern [27]. Nevertheless, these studies had small sample sizes and did not include children with different lesion types.

The aim of this study is twofold: we first investigated the added value of AOT to CIMT in improving UL sensorimotor function in children with uCP; and secondly, we explored the influence of behavioural and neurological factors treatment response.

## 2. Materials and Methods

### 2.1. Participants

This randomized, evaluator-blinded, controlled clinical trial has been fully described elsewhere [31], and will be briefly summarized here. The study was conducted at KU Leuven and was approved by the Ethics Committee of the University Hospitals Leuven (S56513) and registered at www.clinicaltrials.gov (identifier NCT03256357). All children agreed to participate, and their parents or caregivers signed the informed consent.

### 2.2. Study population and randomization

Children with uCP were recruited between June 2014 and June 2017 via the CP reference centre of the University Hospitals Leuven. Inclusion criteria were (1) confirmed diagnosis of uCP, (2) aged 6-12 years, (3) sufficient cooperation to complete the activities and assessments, and (4) minimal ability to actively grasp and stabilize an object with the more impaired hand (House Functional Classification Score (HFC) ≥ 4 [32]). Children were excluded in case of UL surgery in the last 2 years, or botulinum toxin A-injections 6 months prior to enrolment. The participants were stratified according to the HFC scale (4–5 vs. 6–7), age (6-9y vs. 10–12 y), and the CST wiring pattern (contralateral, bilateral, ipsilateral) and assigned to the CIMT+AOT or CIMT+placebo AOT group by using a permuted block design of two. Randomization was performed by a researcher independent of the recruitment and evaluation sessions (HF).

### 2.3. Intervention

The intervention was delivered in a day camp model during 9 out of 11 consecutive days (6 hours/day, total of 54 hours of therapy). During the camp, all children wore a tailor-made hand splint on the less impaired hand while performing unimanual exercises during individual therapy (9h), group activities (30h), and AOT/placebo condition (15h).

The **individual therapy** was based on motor learning principles of shaping and repetitive practice by focusing on four goals: (1) active wrist and elbow extension, (2) forearm supination, (3) grip strength, and (4) fine motor tasks. The **group activities** consisted of painting, crafting, cooking, and outdoor playing, selected to stimulate the intensive use of the more impaired hand. Children assigned to the CIMT+AOT group received 15 hours of **AOT sessions**. During these sessions, children watched video sequences showing unimanual goal-directed actions, adapted to the child’s functional level (Additional files of [31]). The CIMT+placebo group watched video games free of human motion. Afterwards, they executed the same actions for 3 minutes in the same order as the experimental group, for which only verbal, non-suggestive instructions were provided. To account for the compliance of the video observation in the experimental group, a yes/no question related to each video activity was asked after the second execution of each activity (e.g. is the box taken from the top? Did you see the palm of the hand?). At the end of the intervention, the number of correct answers were summed, ranging from 0 (all answers incorrect) to 45 (all answers correct).

### 2.4. Evaluation

An experienced physiotherapist blinded to group allocation (JH) conducted the evaluations at T0 (baseline, 3-4 months before the intervention), T1 (within 4 days before the intervention), T2 (within 4 days after the intervention), and T3 (6 months after the intervention). At T0, we collected descriptive and clinical characteristics to individually set the child’s therapy goals by experienced physiotherapists. Primary and secondary outcome measures were collected at T1, T2, and T3. At T1, we evaluated sensory function, mirror movements, type of brain lesion, and CST wiring pattern as influencing factors.

#### Outcome measures

The ***primary outcome*** measure was the Assisting Hand Assessment (AHA), which evaluates the spontaneous use of the impaired hand during bimanual activities [33,34]. A certified rater scored the videos, blinded to group-allocation and time-point evaluation. The smallest detectable difference is 5 AHA units [35].

***Secondary outcome*** measures comprised body function and activity measures, following a valid and reliable protocol [36]. At *body function level*, we measured muscle tone, muscle strength, and grip strength. Muscle tone was assessed using the Modified Ashworth Scale (MAS [37]) for the shoulder (adductors and internal rotators), elbow (flexors and pronators), wrist (flexors), and hand (finger flexors and thumb adductors) with a total score ranging 0-28. Muscle strength of the shoulder (flexors, adductors, and abductors), elbow (flexors, extensors, supinators, and pronators), and wrist (flexors and extensors) was evaluated using the 8-point ordinal scale of the Medical Research Council [38] with a total score ranging 0-45. Grip strength was assessed using the mean of three maximum contractions with the Jamar^®^ dynamometer (Sammons Preston, Rolyan, Bolingbrook, IL, USA). Movement quality was evaluated with the Melbourne Assessment 2 (MA2) [39,40], resulting in percentages of four subscales measuring range of motion, accuracy, dexterity, and fluency. The test was scored afterwards by a trained physiotherapist blinded to group-allocation and time-point evaluation.

At *activity level*, we included unimanual movement speed and unimanual and bimanual dexterity. Movement speed (time) was evaluated during six unimanual tasks with the modified version of the Jebsen-Taylor Hand Function Test (JTHFT) [41,42]. The smallest detectable difference for the more affected hand is 70 seconds [43]. Unimanual and bimanual dexterity were evaluated using the Tyneside Pegboard Test, an instrumented 9-hole pegboard test [44]. In the unimanual condition, the child moves the pegs as fast as possible from one board to the other, in both directions, using first the less impaired hand. In the bimanual condition, also in both directions, one hand picks up the pegs, transfers them to the other hand through a hole in a Perspex divider placed between the boards, and the other hand inserts the pegs. The test is electronically timed, and results are outputted using a custom-written software (Institute of Neuroscience, Newcastle University, Newcastle upon Tyne, United Kingdom).

Parents were asked to complete the ABILHAND-Kids and the Children’s Hand-use Experience Questionnaire (CHEQ). The ABILHAND-Kids reliably measures the difficulty experienced by the child in performing 21 daily activities [45]. The CHEQ reliably captures the parent’s perspective of the child’s experience while using the more impaired hand during 29 bimanual activities, resulting in three subscales measuring grip efficacy, time used compared to peers, and experience of feeling bothered [46,47].

#### Influencing factors of treatment response

***Sensory assessments*** comprised measures of exteroception (tactile sense), proprioception (movement sense), two-point discrimination (2PD, Aesthesiometer®) and stereognosis (tactile object identification), which have been shown to be reliable in this population [36]. Tactile and movement sense were classified as normal (score 2), impaired (score 1) or absent (score 0). 2PD was classified according to the minimum width between the two points that the children could discriminate: normal (0-4mm, score 2), or impaired (>4mm, score 1) [48].

Tactile object identification was quantified as the number of objects that the child could recognize (0-6). In addition, a kit of 20 nylon monofilaments (0.04g - 300g) (Jamar® Monofilaments, Sammons Preston, Rolyan, Bolingbrook, IL, USA) was used to reliably determine threshold values for touch sensation [49,50]. Touch sensation was categorized as normal (0.008-0.07g), diminished light touch (0.16-0.4g), diminished protective sensation (0.6-2g), loss of protective sensation (4.19-180g) and untestable (300g), according to the manual.

***Mirror movements*** (MM) were quantitatively assessed with the GriFT device during a squeezing task, following the protocol defined by Jaspers et al (2018) [51]. Before performing the task, we tested the maximum voluntary contraction of each hand, starting with the less affected hand. We instructed the children to play a game requiring rhythmic squeezing of one handle with one hand (active hand), while holding the second handle with the other hand (passive hand). The game consisted of controlling with the active hand the position of an astronaut (higher forces corresponding to a higher position on the screen), with the goal to jump over meteorites flying across the screen. MM characterization was based on the comparison of grip force profiles of the active vs. the passive hand, and consisted of the calculation of MM amplitude. MM frequency represents the number of squeezes in the passive hand that exceeded a predefined threshold, expressed as a percentage of the total number of squeezes produced in the active hand. MM amplitude is the average amplitude ratio of the squeezes between both hands, based on only those squeezes in the passive hand that exceeded a predefined threshold. Lastly, MM amount was computed as the frequency by amplitude product, providing an overall indication of the MM occurrence. Full details on the calculation can be found in [51]. MM amount in each hand was used for further statistical analysis.

#### Brain imaging and neurophysiology

The MRI was acquired with a 3T system (Achieva, Philips Medical Systems, Best, The Netherlands) equipped with a 32 channels coil. Structural images were acquired using three-dimensional fluid-attenuated inversion recovery (3D FLAIR) and magnetization prepared rapid gradient echo (MPRAGE). MRI results were used to characterize the lesion type according to the presumed timing (malformation, predominantly white matter, predominantly grey matter) [52] by a child neurologist (EO). To identify the underlying CST wiring pattern, a single-pulse TMS session was conducted. A MagStim 200 Stimulator (Magstim Ltd., Whitland, Wales, UK) equipped with a focal 70mm figure-eight coil and a Bagnoli electromyography system (Delsys Inc., Natick, MA, USA) was used for data acquisition. After identifying the hotspot and the rest motor thresholds, motor evoked potentials were elicited and recorded on the thumb adductor and opponent muscles on both hands, to identify the wiring pattern (contralateral, bilateral or ipsilateral). Children with contraindications to MRI or Transcranial Magnetic Stimulation (TMS) did not undergo the respective measurement. There were no adverse events.

### 2.5 Statistical analyses

#### Effect of the intervention over time

Normality was checked using the Shapiro-Wilk test and inspection of the histograms for symmetry. To conduct parametric statistics, a logarithmic transformation was applied to the parameters of grip strength, the JTHFT, the instrumented pegboard test, the ‘range of motion’ subscale of the MA2, and the ‘feeling bothered’ subscale of the CHEQ questionnaire. A reflect and square root transformation was applied to the ‘accuracy’ subscale of the MA2. Descriptive statistics were reported according to the nature of the data (i.e. means and standard deviations for continuous data and median and interquartile ranges for ordinal data). Mixed models were used to study changes after the intervention over time. By using random effects, these models can correct for the dependency among repeated observations. Furthermore, these models deal with missing data offering valid inferences, assuming that missing observations are unrelated to unobserved outcomes [52]. Changes over time between groups were tested by including group*time interactions. In case of a significant interaction, group-dependent changes were investigated separately in each group. Significant time trends were further inspected using pairwise post-hoc comparisons between T1-T2, T1-T3, and T2-T3 (Bonferroni corrected). The effect sizes of these comparisons were calculated and interpreted according to Cohen’s d formula (small, 0.2–0.5; medium, 0.5–0.8, and large>0.8) [53].

#### Influencing factors of treatment response

Both behavioural (age, initial motor function based on AHA and JTHFT score at T1, sensory function, and MM amount) and neurological characteristics (type of brain lesion and CST wiring pattern) were evaluated as potential influencing factors of treatment response. All variables were included in their original scale except for a dichotomized score for initial motor function. Initial low motor function was defined as either <50 in the AHA units or >350 seconds in the JTHFT (defined as the 25^th^ percentiles for the total group at T1). These variables were included as covariates in the models to influence outcome in the AHA (bimanual) and JTHFT (unimanual), together with the time*group interaction. If the interaction with group was not significant, the interaction with time was examined. Post-hoc analyses with Bonferroni correction were conducted in case of significant interactions and trends (<0.10), as this would allow us to capture tendencies immediately after the intervention.

All statistical analyses were performed using SPSS Statistics for Windows version 25.0 (IBM Corp. Armonk, NY: IBM Corp.). The two-sided 5% level of significance was used for interactions and main effects.

## 3. Results

### 3.1. Participants

Forty-four children participated in the study (mean age (SD) 9y6m (1y10m); 27 boys; 23 left-sided uCP; 9 Manual Ability Classification System I (MACS, [54]), 15 MACS II, and 20 MACS III), and were allocated to the CIMT+AOT group (n=22) and CIMT+placebo group (n=22) (Table 1; Supporting information Table S1). All children completed the intervention program (100% compliance), but two allocated to the CIMT+placebo group were lost to follow-up (Figure 1).

**Table 1.** Demographic characteristics of the participants per group.

|  |  | CIMT+placebo group<br>(n=22) | CIMT+AOT group<br>(n=22) | p-value |
| --- | --- | --- | --- | --- |
| **Age** | mean (SD) | 9y6m (1y10m) | 9y6m (1y11m) | 0.89<sup>1</sup> |
| **Sex** | n (%) |  |  |  |
| Boys |  | 12 (55) | 15 (68) | 0.35<sup>2</sup> |
| Girls |  | 10 (45) | 7 (32) |  |
| **More affected side** | n (%) |  |  |  |
| Left |  | 14 (64) | 9 (41) | 0.13<sup>2</sup> |
| Right |  | 8 (36) | 13 (59) |  |
| **MACS** | n (%) |  |  |  |
| I |  | 3 (14) | 6 (27) | 0.39<sup>2</sup> |
| II |  | 7 (32) | 8 (36.5) |  |
| III |  | 12 (55) | 8 (36.5) |  |
| **HFC system** | n (%) |  |  |  |
| Levels 4-5 |  | 18 (82) | 16 (73) | 0.47<sup>2</sup> |
| Level 6-8 |  | 4 (18) | 6 (27) |  |
| **Lesion type** | n (%) |  |  |  |
| Malformation |  | 0 (0) | 1 (4.5) | 0.18<sup>2</sup> |
| PV lesion |  | 5 (23) | 12 (54.5) |  |
| CSC lesion |  | 13 (59) | 7 (32) |  |
| Acquired |  | 1 (4.5) | 0 (0) |  |
| Other |  | 3 (13.5)<sup>§</sup> | 0 (0) |  |
| Unknown |  | 0 (0) | 2 (9)<sup>°</sup> |  |
| **CST wiring** | n (%) |  |  | 0.42<sup>2</sup> |
| Contralateral |  | 1 (5) | 3 (14) |  |
| Bilateral |  | 8 (36) | 5 (23) |  |
| Ipsilateral |  | 7 (32) | 8 (36) |  |
| Unknown<sup>‡</sup> |  | 6 (27) | 6 (27) |  |

MACS, Manual Ability Classification System; HFC, House Functional Classification; PV, periventricular; CSC, cortico-subcortical; CST, corticospinal tract. <sup>§</sup>Other: brainstem tumor ( $n=1$ ), hemispherectomy ( $n=2$ ); <sup>°</sup>No MRI performed ( $n=1$ panic attack, $n=1$ refused to complete MRI). <sup>‡</sup>TMS not performed or inconclusive. <sup>1</sup>Independent samples t-test, <sup>2</sup>Pearson Chi-Square test.

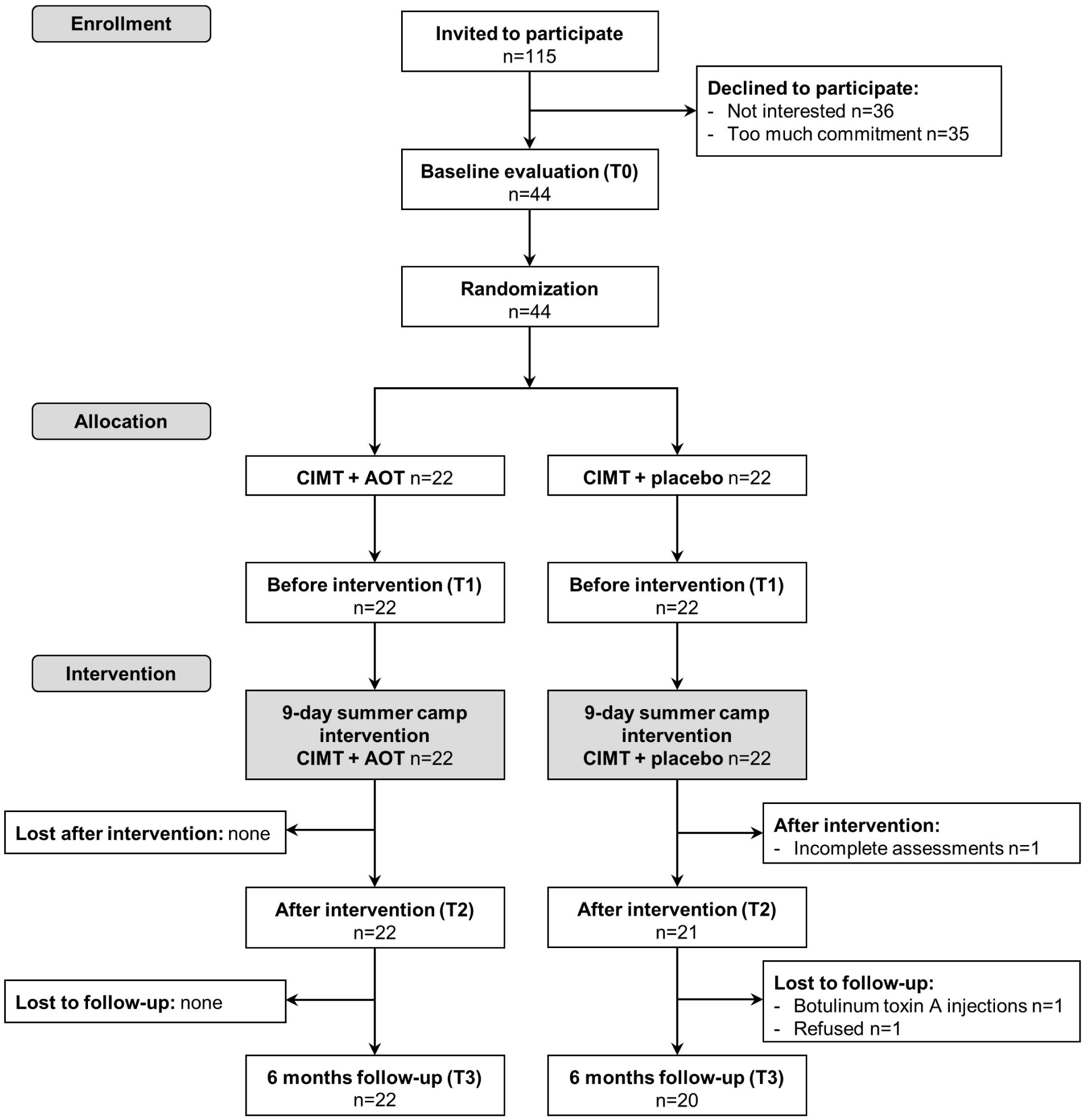

### 3.1. Treatment efficacy

Table 2 summarizes the outcome measures for each intervention group at every time point. All children who received AOT sessions showed a good compliance to the video observation, based on the number of correct answers to the video-related questions (median=42, interquartile range=5, range 30-45). No differences in hand function between groups were found at T1 (all p>0.05, Table S2 Supporting information).

**Table 2.** Estimated marginal means (standard error) of outcome measures at each time point, and statistical comparison (F (p-values)).

|  |  | Total group (n=44) |  |  |  |  |  |  |
| --- | --- | --- | --- | --- | --- | --- | --- | --- |
|  |  | T1 (pre) | T2 (post) | T3 (6m follow-up) | Time*Group | Time | ES T1 vs. T2 | ES T1 vs. T3 |
| **Primary outcome measure** |  |  |  |  |  |  |  |  |
| AHA (units 0-100) | CIMT + placebo | 54.73 (2.64) | 56.59 (2.60) | 56.10 (2.75) | 0.56 (0.58) | **18.62 (<0.001)<sup>SY</sup>** | 0.18 | 0.14 |
|  | CIMT + AOT | 58.68 (2.64) | 61.23 (2.60) | 60.73 (2.75) |  |  |  |  |
| **Secondary outcome measures** |  |  |  |  |  |  |  |  |
| Spasticity (MAS) | CIMT + placebo | 4.60 (0.37) | 4.41 (0.34) | 4.89 (0.38) | 1.56 (0.22) | 2.95 (0.06) | 0.14 | 0 |
|  | CIMT + AOT | 5.05 (0.38) | 4.77 (0.34) | 4.77 (0.37) |  |  |  |  |
| Muscle strength (MMT) | CIMT + placebo | 10.21 (0.20) | 11.11 (0.16) | 10.82 (0.16) | 3.01 (0.06) | **52.07 (<0.001)<sup>S</sup>** | 0.93 | 0.79 |
|  | CIMT + AOT | 10.23 (0.20) | 10.86 (0.16) | 10.89 (0.15) |  |  |  |  |
| Grip strength (Kg) | CIMT + placebo | 4.06 (1.15) | 5.24 (1.13) | 6.10 (1.13) | 0.49 (0.62) | **16.44 (<0.001)<sup>SY</sup>** | 0.16 | 0.24 |
|  | CIMT + AOT | 4.47 (1.16) | 5.58 (1.13) | 5.94 (1.13) |  |  |  |  |
| MA2 – Range of Motion (%) | CIMT + placebo | 62.57 (1.05) | 65.09 (1.05) | 68.08 (1.06) | 2.95 (0.06) | **3.51 (0.04)<sup>Y</sup>** | 0.13 | 0.44 |
|  | CIMT + AOT | 68.61 (1.05) | 67.65 (1.05) | 69.12 (1.06) |  |  |  |  |
| MA2 – Dexterity (%) | CIMT + placebo | 59.07 (4.02) | 62.96 (3.83) | 62.56 (4.29) | 1.18 (0.32) | 2.12 (0.13) | 0.10 | 0.12 |
|  | CIMT + AOT | 68.80 (4.03) | 68.73 (3.81) | 70.06 (4.26) |  |  |  |  |
| MA2 – Accuracy (%) | CIMT + placebo | 88.33 (0.81) | 87.17 (0.86) | 91.92 (0.85) | 2.10 (0.14) | 0.51 (0.61) | 0 | 0 |
|  | CIMT + AOT | 94.42 (0.81) | 94.91 (0.86) | 93.74 (0.86) |  |  |  |  |
| MA2 – Fluency (%) | CIMT + placebo | 78.36 (2.80) | 77.27 (2.61) | 77.71 (2.71) | 2.08 (0.14) | 2.92 (0.07) | 0.15 | 0.06 |
|  | CIMT + AOT | 82.60 (2.82) | 80.09 (2.60) | 84.71 (2.68) |  |  |  |  |
| JTHFT (s) | CIMT + placebo | 211.94 (1.17) | 155.83 (1.18) | 176.94 (1.18) | 3.21 (0.05) | **26.40 (<0.001)<sup>SY</sup>** | 5.81 | 5.21 |
|  | CIMT + AOT | 172.29 (1.17) | 140.42 (1.18) | 132.56 (1.18) |  |  |  |  |
| IPT – Large pegs bimanual (more aff toward less-aff) | CIMT + placebo | 39.65 (1.10) | 30.45 (1.11) | 32.65 (1.10) | 1.90 (0.16) | **20.98 (<0.001)<sup>SY</sup>** | 0.85 | 0.80 |
|  | CIMT + AOT | 35.83 (1.10) | 32.73 (1.10) | 31.30 (1.10) |  |  |  |  |
| IPT - Large pegs bimanual (less-aff toward more-aff) | CIMT + placebo | 61.58 (1.16) | 48.76 (1.16) | 51.73 (1.16) | 0.15 (0.86) | **6.73 (0.004)<sup>SY</sup>** | 1.44 | 1.20 |
|  | CIMT + AOT | 53.17 (1.15) | 44.64 (1.15) | 45.51 (1.15) |  |  |  |  |
| IPT – Large pegs unimanual (more-aff hand) | CIMT + placebo | 44.90 (1.14) | 37.17 (1.12) | 37.87 (1.12) | 0.13 (0.88) | **7.46 (0.002)<sup>SY</sup>** | 0.93 | 0.93 |
|  | CIMT + AOT | 38.13 (1.14) | 32.30 (1.11) | 31.96 (1.11) |  |  |  |  |
| IPT – Medium pegs unimanual (more-aff hand) | CIMT + placebo | 42.15 (1.13) | 37.40 (1.12) | 36.71 (1.14) | 0.01 (0.99) | **7.40 (0.002)<sup>SY</sup>** | 0.65 | 0.77 |
|  | CIMT + AOT | 39.62 (1.12) | 35.02 (1.11) | 34.07 (1.13) |  |  |  |  |
| IPT – Small pegs unimanual (more-aff hand) | CIMT + placebo | 73.44 (1.17) | 55.07 (1.16) | 56.97 (1.17) | 0.80 (0.46) | **13.13 (<0.001)<sup>SY</sup>** | 1.57 | 1.63 |
|  | CIMT + AOT | 50.33 (1.16) | 43.76 (1.15) | 41.39 (1.16) |  |  |  |  |

**Table 2 (continuation).**
|  |  | Total group (n=44) |  |  |  |  |  |  |
| --- | --- | --- | --- | --- | --- | --- | --- | --- |
|  |  | T1 (pre) | T2 (post) | T3 (6m follow-up) | Time*Group<br>p | Time | ES T1 vs.<br>T2 | ES T1 vs.<br>T3 |
| ABILHAND-Kids (logits) | CIMT + placebo | 1.64 (0.25) | 1.90 (0.66) | 1.79 (0.28) | 1.04 (0.37) | 0.44 (0.65) | 0.08 | 0.11 |
|  | CIMT + AOT | 1.67 (0.25) | 1.63 (0.26) | 1.77 (0.28) |  |  |  |  |
| CHEQ – Grip efficacy (%) | CIMT + placebo | 45.78 (3.20) | 48.15 (2.78) | 49.30 (3.73) | 0.40 (0.68) | 3.24 (0.05) | 0.30 | 0.30 |
|  | CIMT + AOT | 42.08 (3.24) | 47.86 (2.64) | 47.75 (3.43) |  |  |  |  |
| CHEQ – Time used (%) | CIMT + placebo | 39.15 (3.18) | 44.69 (2.99) | 41.62 (3.21) | 0.45 (0.64) | **5.38 (0.009)<sup>§</sup>** | 0.31 | 0.22 |
|  | CIMT + AOT | 38.17 (3.23) | 41.86 (2.93) | 41.82 (2.99) |  |  |  |  |
| CHEQ – Feeling bothered (%) | CIMT + placebo | 40.02 (1.10) | 50.03 (1.06) | 46.24 (1.06) | 3.12 (0.06) | **3.66 (0.04)<sup>§</sup>** | 0.85 | 0.78 |
|  | CIMT + AOT | 45.03 (1.10) | 47.74 (1.06) | 49.20 (1.06) |  |  |  |  |

AHA, Assisting Hand Assessment; MAS, Modified Ashworth Scale; MMT, Manual Muscle Testing; Kg, Kilograms; MA2, Melbourne Assessment 2; JTHFT, Jebsen-Taylor Hand Function test; IPT, Instrumented Pegboard Test; CHEQ, Child's Hand Experience Questionnaire; T1, pre-intervention evaluation; T2, post-intervention evaluation; T3, 6 months follow-up evaluation; CIMT, modified Constraint Induced Movement Therapy; AOT, Action-Observation Training. <sup>§</sup>Significant at the T1 vs. T2 comparison. <sup>¥</sup>Significant at the T1 vs. T3 comparison.

#### The added value of AOT to CIMT

We did not find between-groups differences in improvements over time on the primary outcome (AHA; p>0.05) nor on the secondary outcomes (all p>0.05, Table 2).

#### Improvements over time

##### Primary outcome

The descriptive data is shown in Table S3 of Supporting information. The total group improved over time in the AHA (p<0.001), with a significant mean improvement of 2.21 AHA units immediately after the intervention (T1-T2, p<0.001) and maintained at follow-up (T2-T3, p<0.001). Effect sizes were low (ES=0.14-0.18). Immediately after the intervention, most of the children (n=32, 74%) improved their AHA score, of which 28% (n=9) showed an improvement ≥ 5 AHA units (Figure 2A).

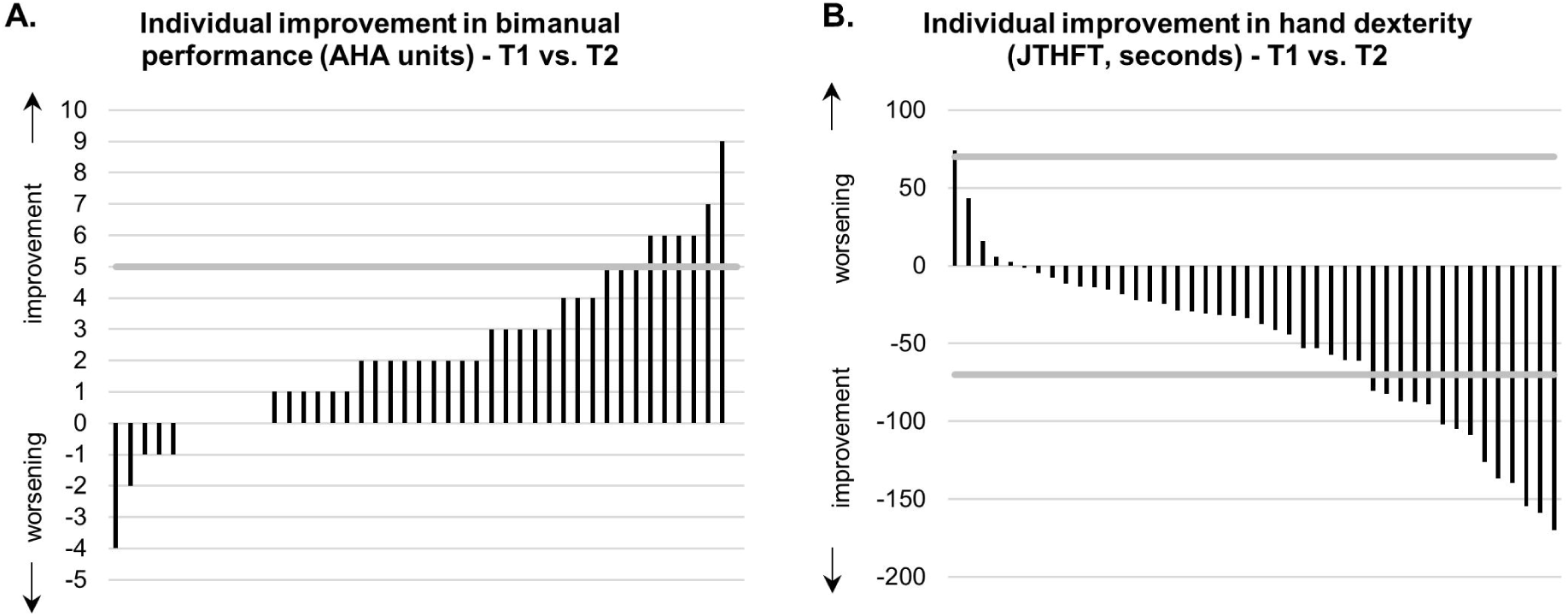

##### Secondary outcomes

At <u>body function level</u>, we found an improvement in grip and muscle strength (p<0.001), occurring immediately after the intervention (p<0.001) and maintained at follow-up (p<0.001). From the MA2 scale, only range of motion improved over time (p=0.04), although the improvements were not immediately after the camp (p>0.05), but at follow-up (p=0.04). No significant changes were found for spasticity scores (p>0.05).

At <u>activity level</u>, we found large improvements in movement speed (JTHFT, p<0.001), performing on average 43 seconds faster after the intervention (p<0.001) and retaining the gains at follow-up (p<0.001). After the intervention, 88% of the children (n=37) improved, and 32% (n=14) improved more than the smallest detectable difference (70 seconds) (Figure 1B) of which 70% (n=31) maintained these gains at follow-up. The improvements were also large in unimanual and bimanual dexterity (pegboard test, all p<0.02). Unimanual dexterity improved immediately after the intervention (p<0.01) and improvements were retained at follow-up (p<0.05). The ‘small pegs’ condition was incomplete for 8 children before the camp, although 6 of these 8 children could complete the task after the camp, and 4 of them still completed it at follow-up. Bimanual dexterity also improved immediately after the intervention (p<0.01) and at follow-up (p<0.01). Lastly, the CHEQ results showed a reduction in time consumption and feeling bothered while performing activities (p=0.009 and p=0.04, respectively), increasing by 4.47% (T1 vs. T2, p=0.008) and 5.96% (T1 vs. T2, p=0.03), respectively. ABILHAND-Kids did not change after the intervention (p=0.65).

In summary, we found improvements after the intervention on muscle strength (ES 0.93) and grip strength (ES 0.16), on unimanual dexterity measured with the JTHFT (ES 5.81) and in unimanual (ES 0.65-1.57) and bimanual dexterity (ES 0.85-1.44) measured with the instrumented pegboard test. Lastly, the subscales of the CHEQ ‘feeling bothered’ and ‘time used’ improved with large (ES 0.85) and small (ES 0.31) effect size, respectively. Additionally, the retained gains were also captured by these assessments with similar ES.

### 3.2. Influencing factors

We evaluated the influence of behavioural and neurological characteristics on treatment outcome for the primary outcome measure (AHA) and for movement speed (JTHFT), as it showed the largest effect size (>5, Table 2). An overview of the statistical results is reported in Table 3. Table S4 in “Supporting information” reports the number of children included in each category for the significant influencing factors.

**Table 3.**
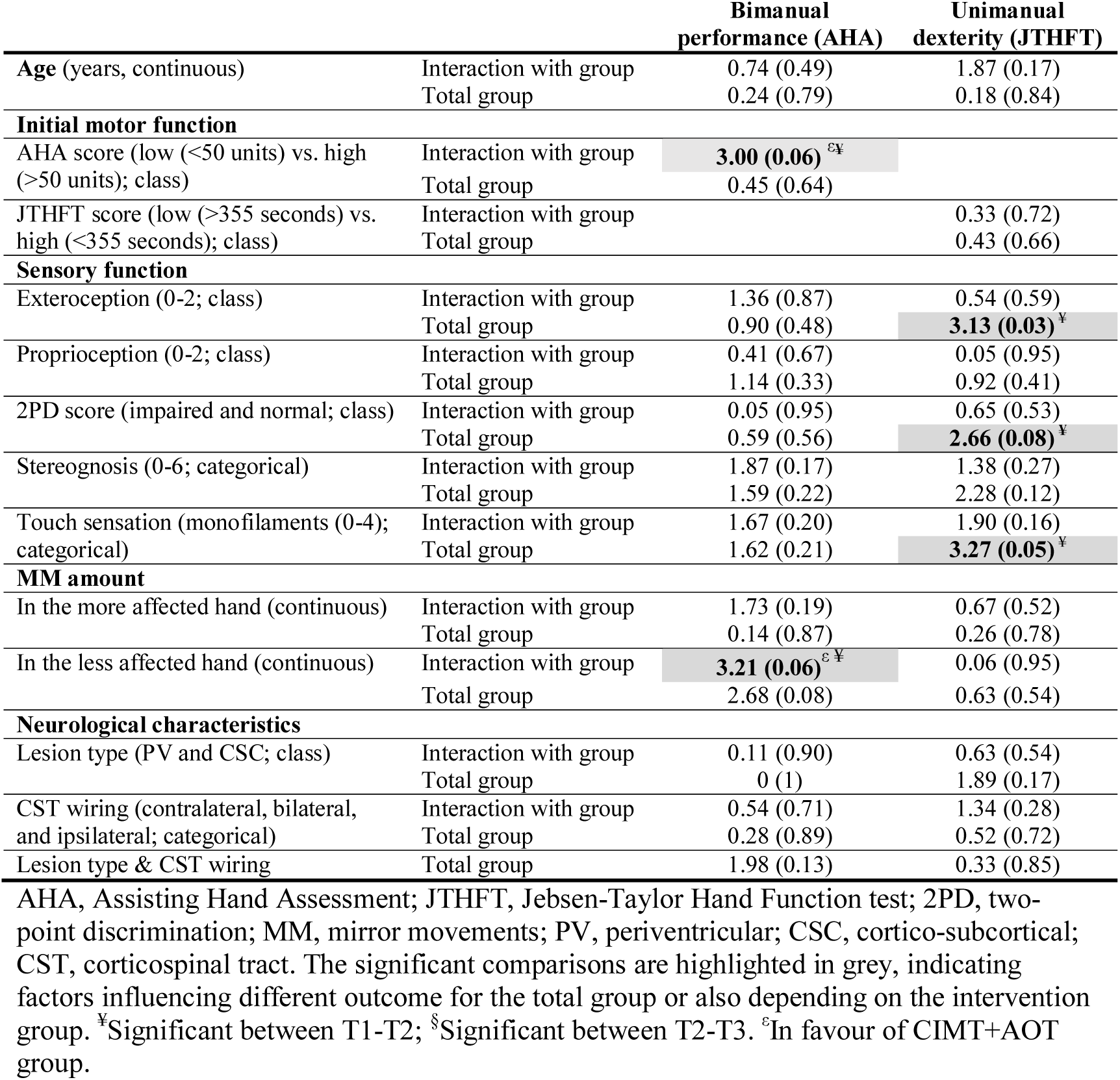
Statistical inference overview of the influencing value of behavioural, and neurological characteristics on treatment response (F (p-values)).

#### Are there subgroups of children who respond better to AOT?

We found a trend toward a significant influence of **initial hand function** (AHA score) on treatment response at three time points (F=3.00, p=0.06; Figure 3A), which was significant between T1-T2 (p=0.02). This interaction indicated that if the initial AHA score was low, the CIMT+AOT group benefitted more than the CIMT+placebo group. If the initial AHA score was high, both groups improved equally (Figure 3B).

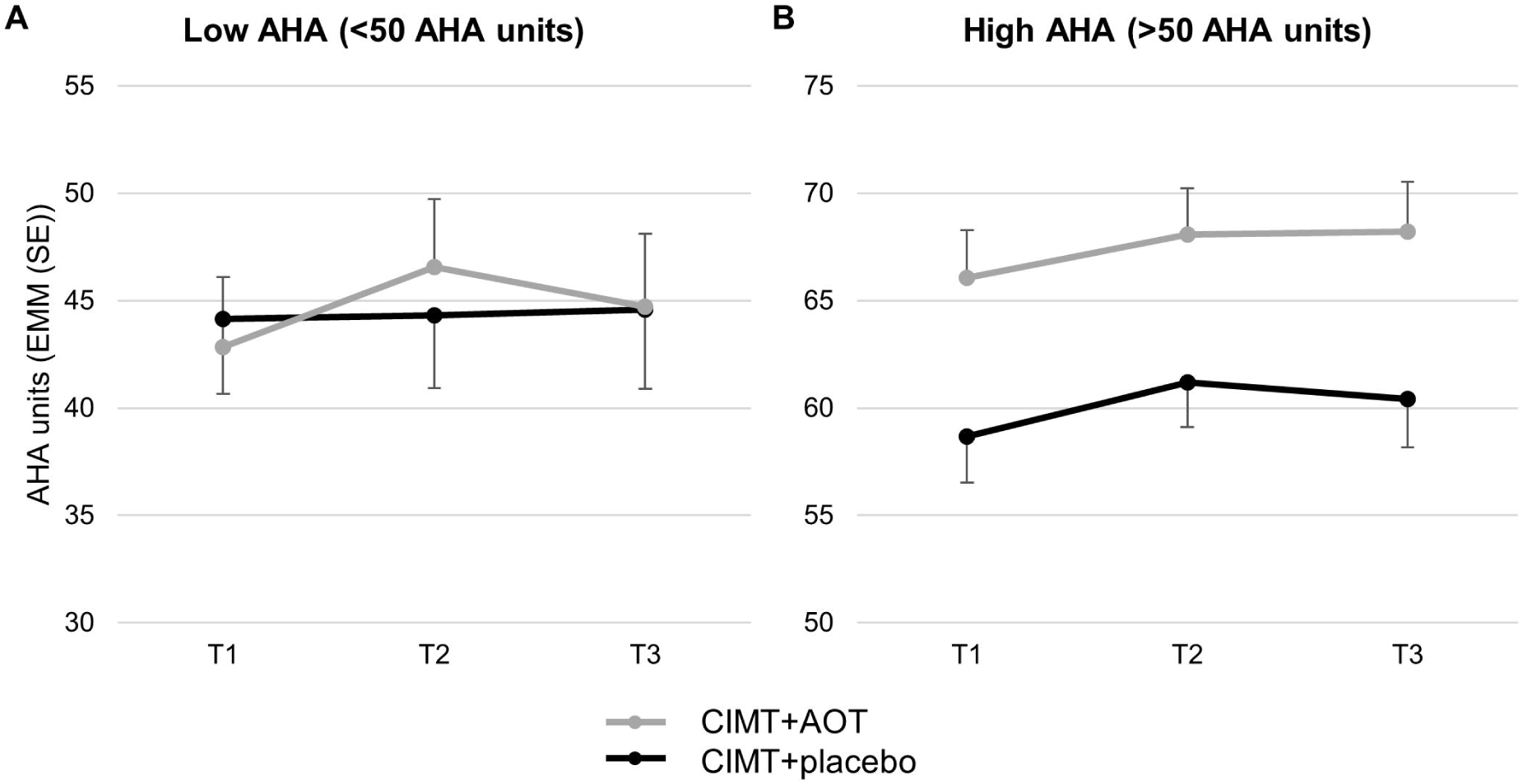

Similarly, we found a trend toward a significant influence of **MM amount** in the less affected hand (more affected hand actively moving) on treatment response of bimanual performance (F=3.21, p=0.06, Figure 4). This interaction indicated that if the initial amount of MM was high, the CIMT+AOT group benefitted more than the CIMT+placebo group. If the initial MM amount was low, both groups improved similarly.

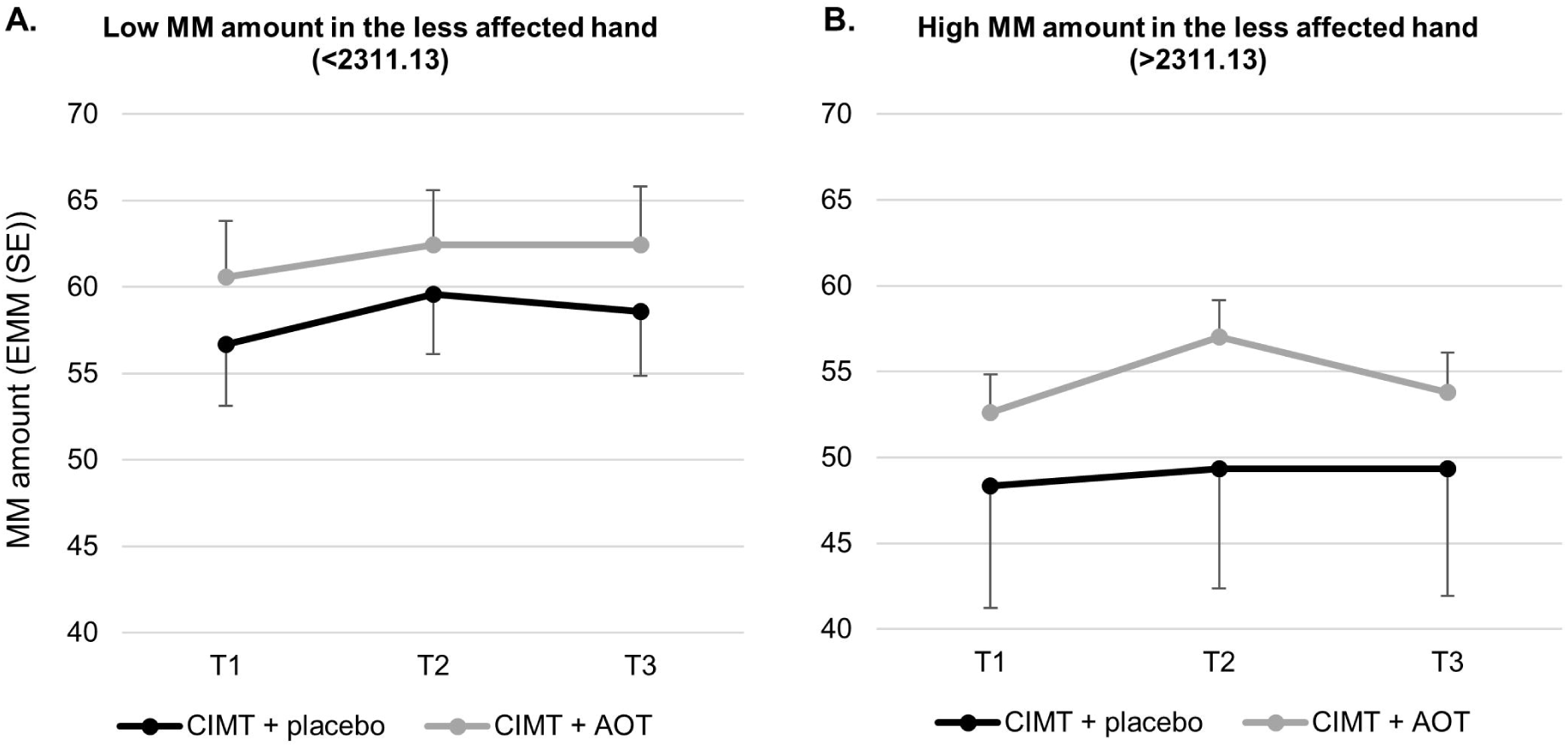

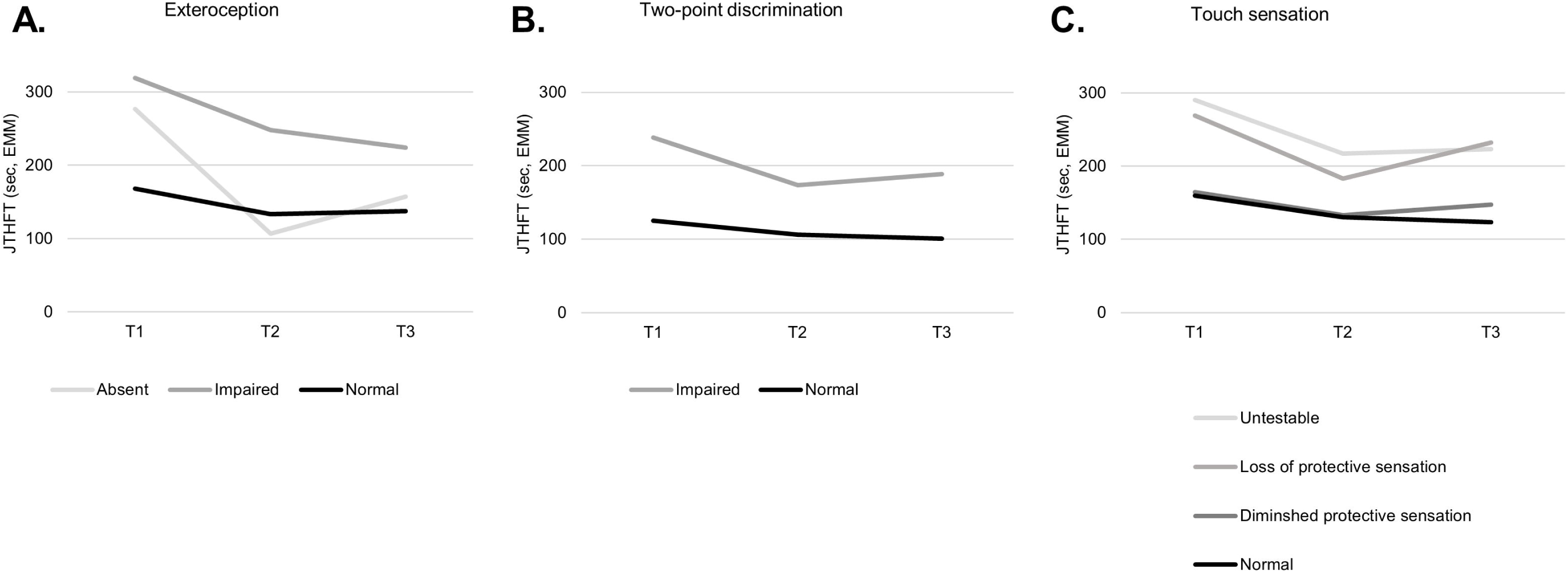

Responsiveness to AOT did not depend on age nor sensory function for either AHA or JTHFT (p>0.05). Regarding neurological characteristics, neither type of brain lesion nor CST wiring pattern had an influence on responsiveness to AOT (p>0.05).

#### Are there subgroups of children who respond better to CIMT with or without AOT?

We found that sensory function was able to influence treatment response for the total group for unimanual dexterity. More specifically, exteroception, 2PD, and touch sensation influenced the outcome of the JTHFT (p=0.03-0.08; Table 3), indicating that children with more impaired sensory function benefitted more from the CIMT intervention, compared to those with normal sensory function (Figure 4). Note that initial motor function did not interfere with these interactions (interaction term p<0.05).

The responsiveness to CIMT with or without AOT did not depend on age, initial motor function, stereognosis, nor amount of MM (p>0.05, Table 3). Similarly, neither type of brain lesion nor CST wiring pattern had an influence on responsiveness to CIMT with or without AOT when tested individually (p>0.05, Figure 6) or combined (interaction between CST wiring pattern and type of the brain lesion, p>0.05).

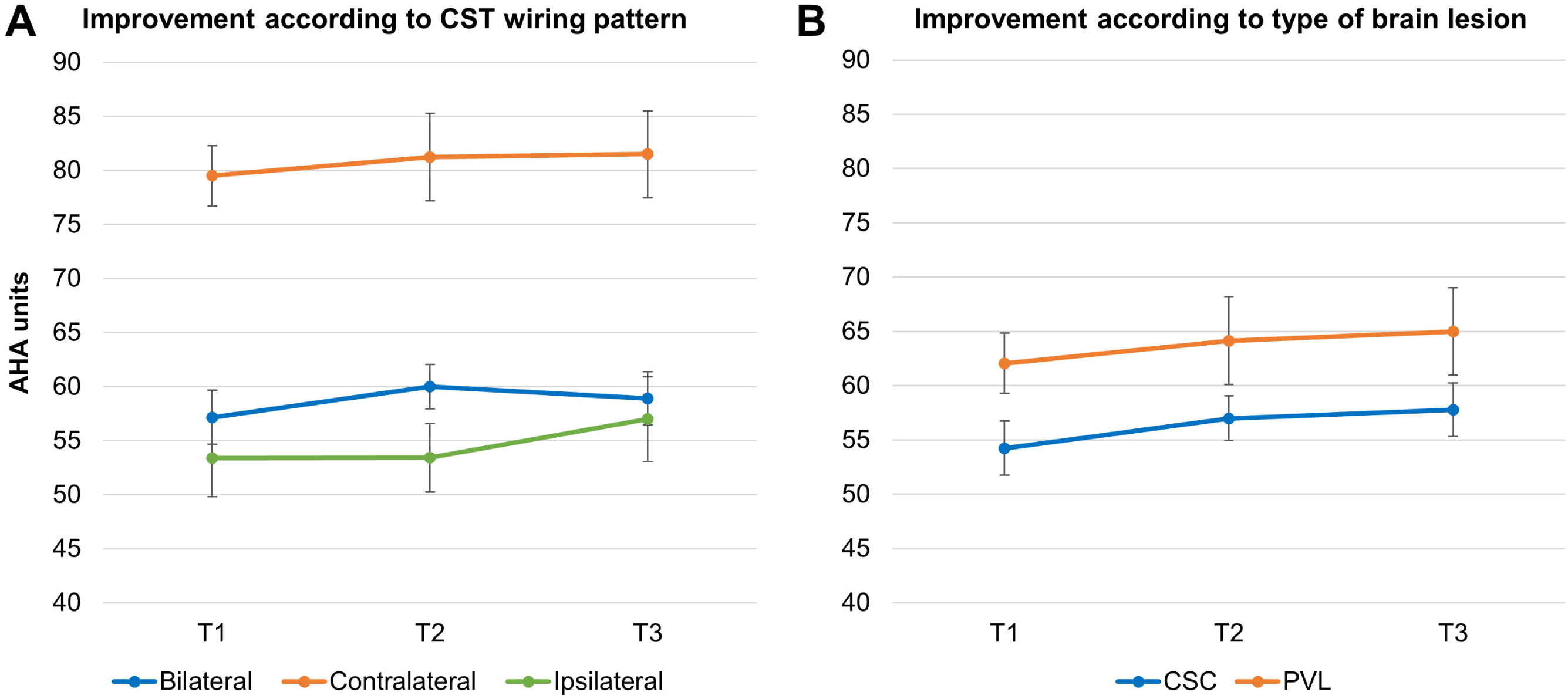

## 4. Discussion

This randomized controlled trial was the first to investigate the effects of an intensive camp-based treatment model combining CIMT and AOT to improve UL function at body function and activity level, by including clinical and instrumented outcome measures, as well as both behavioural and neurological factors to determine their influence on treatment outcome. The effects showed that, with or without AOT, an intensive CIMT training approach delivered in a summer camp setting leads to improvements in UL function. Although we could not demonstrate an overall added effect of AOT, our results suggest that the addition of AOT to CIMT may be beneficial for children with initial poor hand function and high amount of MM. In addition, we found that sensory function influenced treatment response following CIMT (with or without AOT).

### What is the added value of AOT to CIMT on UL function?

The novelty of this RCT lies in the investigation of the added value of AOT to a well-established therapy approach, such as CIMT. Overall, our results show similar improvements in the CIMT+AOT and CIMT+placebo group. To date, the first studies exploring AOT in children with uCP have proven its effectiveness in improving UL motor function [9–12]. However, these studies investigated the effect of AOT alone compared to a placebo or no observation, and not the added effect of AOT to a well-established UL therapy, such as CIMT. Our results are in agreement to Kirkpatrick et al (2016), who found no effect of AOT compared to repetitive practice in a home setting [13]. In our study, the lack of additional value of AOT could be explained by the large gains after CIMT reported in current and previous studies [14,55,56].

Interestingly, we identified that children with initial poorer hand function (lower than 50 AHA units) benefitted more from the combined approach CIMT+AOT compared to CIMT alone. On average, the group receiving CIMT+AOT with initially poorer AHA scores improved 4 AHA units, while the CIMT+placebo group did not improve. Sgandurra et al (2018) recently reported that poorer bimanual performance, measured with the AHA, was indicative for a more lateralized mirror neuron system in children with uCP (toward the non-lesioned hemisphere) [57]. Given these results, it makes sense that the additional AOT intervention for children who had poorer bimanual performance was more effective, as AOT may have facilitated the activation of their mirror neuron system through the video-observation. For those children who showed high bimanual performance, it seems plausible that the mirror neuron system cannot be further facilitated, as this may be intact, and other sensorimotor brain regions would need to be stimulated to further increase their motor function, for example to facilitate inter-hemisphere connectivity. Also, in contrast to other studies, the current study only included unimanual tasks to fit the unimanual concept of CIMT, although more challenging bimanual AOT tasks may be needed to further improve UL function in children with initially better hand function. While this finding is clinically relevant, further studies are clearly needed to confirm our results, as well as to define the best delivery of AOT.

A second significant influencing factor of treatment outcome between groups following the intervention was *amount of MM* in the less affected hand. Whilst children with few MM responded similarly to either training, children with high amount of MM in the less affected hand seemed to improve more after the CIMT+AOT training. There is evidence for a relation between poor bimanual function and high amount of MM [21]. Thus, this result is in line with the previous result, where poor hand function influenced treatment outcome. Unfortunately, we cannot be certain that these changes are not led by concomitant reduction in amount of MM. Additional studies including an evaluation of MM before and after intervention are needed to further clarify these relationships. Nevertheless, this novel finding points toward the importance of measuring MM and it is a first step toward the delineation of training strategies based on clinical characteristics.

### What are the effects of CIMT (with or without AOT) on UL function?

Our results for the total group showed improvements in grip and muscle strength (ES 0.16-0.93), JTHFT (ES 5.81) and the instrumented pegboard test (ES 0.65-1.57), indicated by their large effect sizes. Moreover, the gains in muscle strength and unimanual dexterity were translated to bimanual dexterity, measured with the bimanual conditions of the instrumented pegboard test (ES 0.65-1.44). Interestingly, these gains also resulted in an increased comfort in using the hand in daily activities as confirmed by the improvement in the CHEQ-subscale ‘feeling bothered’ with large effect size (0.85). Still, this contrasts with the small effect size found for the AHA (0.18). The effect size of the AHA reported in previous CIMT studies in a camp model varies across studies: larger effect sizes (around 1.12) in younger children (18m-8y) [58,59], smaller effects (0.16-0.28) in children aged similar to our study [27,60,61]. According to Hung et al (2011), the AHA measures the quality of the assisting hand while performing bimanual movements and misses the spatiotemporal control of bimanual functioning [62]. Integrating other measures of spatiotemporal control may help to capture these aspects.

In most measures, we found that the improvements were not only seen immediately after the therapy, but also after 6 months, which is in agreement with previous studies [18,27,42,60,61]. This maintenance is of clinical relevance, as intensive therapies are given in shorter periods. Charles et al (2007) showed, however, that between 6 months and 1 year after the first camp, children typically showed a slight functional decline, and a second CIMT dose 1 year after resulted in continued improvements [63]. Boosts of intensive interventions with 6-months or 1-year interval may result in long-lasting and clinically relevant effects.

Interestingly, we also found that children with *impaired sensory function* benefitted more from the intervention compared to children with normal sensory function. To the best of our knowledge, this is the first time that sensory function is investigated as an influencing factor of response to CIMT in children with uCP. The sensory deficits may lead to a failure to use the motor functions and capacities of the more affected UL for spontaneous use in daily life. This phenomenon is known as developmental disregard [64,65]. It is hypothesized that children with developmental disregard may respond better to CIMT due to the forced use of the more affected limb. The positive effects are however partially lost at follow-up, potentially due to the lack of ongoing stimulation of the more affected limb,

### Does the response to CIMT depend on the underlying neurological characteristics?

In our study, we found that all children improved after a CIMT program, irrespective of their lesion type or CST wiring pattern. Interestingly, and adding to the controversy in the literature [27–30], having an ipsilateral CST wiring pattern did not impede improvement after treatment, as these children improved almost 5 AHA units after CIMT (with or without AOT) (see Figure 6). Staudt et al (2014) proposed that one neurological factor is insufficient to impact treatment response after CIMT [66], and a multifactorial model including several neurological characteristics may have larger influence than any factor alone. Nevertheless, our study did not find that the interaction between the lesion type and the CST wiring pattern had a larger influence on treatment outcome. Our study results highlight the variability within each group, suggesting the influence of other factors. We hypothesize that functional measures of both sensory and motor function, and how these functions are integrated in the brain (sensorimotor integration) may be an important influencing factor of treatment outcome. Further investigations including both clinical and neurophysiological measures of the motor and sensory system (motor and sensory evoked potentials), as well as of sensorimotor integration (e.g. with the short latency afferent inhibition protocol [67]) are warranted.

Whilst this study was the first to investigate the merit of AOT in combination with CIMT in a camp model, its limitations should also be addressed. Firstly, we included 44 children according to the power calculation for the primary outcome measure [31]. This sample size could be too low when investigating influencing factors of treatment response, particularly for the neurological characteristics. A second limitation is the lack of a specific outcome measure that evaluates motor planning as targeted with the AOT [11]. Future studies investigating the added effect of AOT should also include such outcome measures, for example the end-posture comfort [68,69]. Lastly, it remains debatable whether a two-week camp can be translated to routine clinical practice, as it demands high commitment from both the parents and the children during the holiday period. In our study, despite a good cooperation during the AOT intervention, the children generally reported that the AOT sessions were monotonous. Hence, we advocate for trainings that are engaging and motivating for the children. For example, a virtual reality environment [70,71] where the child sees himself as an avatar, could serve as a more motivating, engaging, and potentially more effective AOT approach.

In the future, it is crucial that forces between centres and institutions are joined to coordinate multicentre RCTs, which will contribute to fine-tune the identification of responders vs. non-responders through clinical and neurological predictors in a statistically powerful study. Furthermore, future studies should investigate the neuroplastic changes derived from an intensive intervention.

## Conclusion

AOT did not show an overall added effect on improving UL function in children with uCP when combined with CIMT in an intensive training approach. Still, AOT seemed to have an additional positive value in children with poor motor function and high amount of MM, suggesting that the responsiveness to AOT is patient specific. Such insights provide a further step toward patient-tailored intervention approaches. The findings of this study also confirm the efficacy of intensive models of CIMT interventions (with or without AOT), with large effect sizes found in unimanual and bimanual dexterity, which seems to be more beneficial for children with impaired sensory function. The novelty of this study lies within the exploration of behavioural and neurological influencing factors on treatment response, which paves the way toward an effective and individualized treatment planning for children with uCP.

## Data Availability

The data is available upon request to the corresponding author

## 7. Acknowledgments

We would like to express our deepest gratitude to the children and families who participated in this study. We also thank Catherine Huenaerts and Saranda Bekteshi for scoring the video-based assessments, as well as Katrien Fagard for her assistance in the recruitment phase. We are grateful to the master students of physiotherapy who participated in the summer camps and assisted in the measurements. We also thank the schools (Sint Gerardus School in Diepenbeek, DVC Sint-Jozef in Antwerp, VZW Sint-Lodewijk in Wetteren) that let us organize the summer camps in their centres.

